# Viral Pneumonia is Associated with Increased Risk and Earlier Development of Post-Inflammatory Pulmonary Fibrosis

**DOI:** 10.1101/2021.03.08.21252412

**Authors:** Abbas Shojaee, Jonathan M. Siner, Andrey Zinchuk, Yalda Aryan, Naftali Kaminski, Charles S. Dela Cruz

**Affiliations:** Pulmonary, Critical Care and Sleep Medicine Section, Department of Internal Medicine, Yale University School of Medicine, New Haven, Connecticut; Department of Computer Engineering, Faculty of Engineering, Arak University, Arak, Iran

## Abstract

Severe inflammatory response, acute respiratory distress syndrome, and death are established serious consequences of the acute phase of severe viral pneumonia. However, the long-term respiratory outcomes of severe viral pneumonia, including its association with pulmonary fibrosis, are less known.

**Objective:** To determine whether viral pneumonia is associated with an increased incidence of post-inflammatory pulmonary fibrosis.

**Design:** We performed two retrospective observational cohort studies using longitudinal hospitalization records from the States of California (2005-2011) and Florida (2009-2015) for the discovery and validation studies, respectively. Patients who were 85-years-old and younger with at least two hospital encounters but without a prior diagnosis of pulmonary fibrosis were included. International Classification of Diseases-9 (ICD9) codes of primary and secondary diagnoses and procedures were used to identify the **exposure:** diagnosis of viral pneumonia; the **outcome:** incidence of post-inflammatory pulmonary fibrosis [PIPF, ICD9: 515]; and the confounders.

**Methods:** Chronological age was used as the study time scale. Non-parametric Kaplan-Meier (KM) estimator and semiparametric Cox Proportional Hazard modelling were used to assessing the risk of PIPF. P-values < 10^−3^ were considered significant.

**Results:** Among 9,802,565 patients from California and 8,741,345 patients in Florida cohorts, the prevalence of PIPF was 0.61% and 0.62% over 7 and 6.75 years, respectively. Patients with incident PIPF were older than those without [68(SD: 11) vs. 40(22) years]; among patients with PIPF, those with viral pneumonia diagnosis were younger than those without [63(12) versus 68(11) years]. Incidence of PIPF was higher for those with viral pneumonia diagnosis versus those without [1.6 (CI:1.51-1.69) vs. 0.91 (CI:0.86-0.96)] cases per 1000 person-years in California and in Florida [1.11 (CI:1.06 −1.16) vs, 0.93 (CI:0.89-0.98)]. Viral pneumonia was associated with increased risk of incident PIPF in both California aHR = 1.49 (1.38, 1.61), and Florida aHR of 1.26 (1.20, 1.33) cohorts (Table). Among patients who developed PIPF, the median time to diagnosis was 7.41 (6.16 −8.66) and 4.80 (4.34 - 5.26) years earlier for patients with viral pneumonia versus without in California and Florida cohorts. The association of viral pneumonia was not found for idiopathic pulmonary fibrosis [ICD9: 516.3]. Our findings suggest that patients hospitalized with viral pneumonia may have long term respiratory sequela that is often overlooked and suggest a need for additional studies focusing on phenotyping susceptible patients. This finding is especially important in light of the current COVID-19 pandemic because viral pneumonia is the most common manifestations of the disease, which could lead to subsequent fibrosis.

## Background

Severe inflammatory response, acute respiratory distress syndrome (ARDS), and death are established serious consequences of the acute phase of severe viral pneumonia. However, the long-term respiratory outcomes of severe viral pneumonia, including its association with pulmonary fibrosis, are less known.

### Objective

To determine whether viral pneumonia is associated with an increased incidence of post-inflammatory pulmonary fibrosis.

### Design

We performed two retrospective observational cohort studies using longitudinal hospitalization records from the States of California (2005-2011) and Florida (2009-2015) for the discovery and validation studies, respectively.

### Data

Administrative health data from Healthcare Research and Quality Healthcare Cost and Utilization Project’s(HCUP) State Inpatient Database (SID) and State Emergency Department Database (SEDD) (1, 2). The SID data capture over 97% of all hospitalization discharges in the U.S. and SEDD files capture emergency department visits that did not result in hospitalization.(3-5). Each record captures all procedures and diagnosis codes using the International Classification of Diseases, Ninth Edition, Clinical Modification (ICD-9CM), as well as demographics, payment, and length-of-stay for each patient.

### Participants

Patients who were 85-years-old and younger with at least two hospital encounters but without a prior diagnosis of pulmonary fibrosis were included. International Classification of Diseases-9 (ICD9) codes of primary and secondary diagnoses and procedures were used to identify the **exposure:** diagnosis of viral pneumonia; the **outcome:** incidence of post-inflammatory pulmonary fibrosis [PIPF, ICD9: 515]; and **confounders:** bacterial pneumonia, sex, asthma, chronic obstructive pulmonary disease, sarcoidosis, rheumatoid arthritis, scleroderma, asbestosis, bronchiectasis, ARDS, mechanical ventilation, smoking, and the time from first to the last observation. History of smoking was identified using the ICD9CM codes including, 305.1 (tobacco use disorder), 649.0 (tobacco use disorder complicating pregnancy, childbirth, or the puerperium), v15.82 (personal history of tobacco use), and 989.84 (toxic effect of tobacco).

## Methods

Chronological age was used as the study time scale (6). Non-parametric Kaplan-Meier (KM) estimator curves were produced and assessed for proportionality of hazard. Semiparametric Cox Proportional Hazard modeling was used to assess the risk of PIPF. Univariate hazard ratio (HR) and HR adjusted for all confounders (aHR) with 95% confidence intervals (CI) are reported. P-values < 10^−3^ were considered significant. Statistical analyses were performed using R software, version 3.5.3 (R Project for Statistical Computing; R Foundation).

## Results

Among 9,802,565 patients from California and 8,741,345 patients in Florida cohorts, the prevalence of PIPF was 0.61% and 0.62% over 7 and 6.75 years, respectively. Patients with incident PIPF were older than those without [68±11 vs. 40±22 years]; among patients with PIPF, those with viral pneumonia diagnosis were younger than those without [63±12 versus 68±11 years]. Incidence of PIPF was higher for those with viral pneumonia diagnosis versus those without [1.6 (CI:1.51-1.69) vs. 0.91 (CI:0.86-0.96)] cases per 1000 person-years in California and in Florida [1.11 (CI:1.06 −1.16) vs, 0.93 (CI:0.89-0.98)]. Viral pneumonia was associated with increased risk of incident PIPF in both California aHR = 1.49 (1.38, 1.61), and Florida aHR of 1.26 (1.20, 1.33) cohorts (Table). Among patients who developed PIPF, the median time to diagnosis was 7.41 (6.16 −8.66) and 4.80 (4.34 − 5.26) years earlier for patients with viral pneumonia versus without in California and Florida cohorts (Figure). The association of viral pneumonia was not found for idiopathic pulmonary fibrosis [ICD9: 516.3] (7).

**Table:** Frequency of Viral Pneumonia and Post Inflammatory Pulmonary Fibrosis and confounders in the California and Florida cohorts, Incidence, Attributable Risk in the population and patients exposed to Viral Pneumonia in California and Florida cohorts are presented along with Risk Ratio, Univariate and multivariable Cox model hazard ratios.

|  | California [No. (%)] |  | Florida [No. (%)] |  |
| --- | --- | --- | --- | --- |
|  | No Viral P. | Viral P. | No Viral P. | Viral P. |
| **Total** | 9,593,937 | 113,847 | 8,498,126 | 244,775 |
| **Age mean (±SD)** | 40.75 (22.25) | 28.83 (21.26) | 40.5 (22.89) | 25.73 (22.21) |
| **Female** | 5,458,446 (56.9) | 65,402 (57.4) | 4,624,168 (54.4) | 136,004 (55.6) |
| **Smoking** | 564,772 (5.9) | 10,593 (9.3) | 348,423(4.1) | 18,191 (7.4) |
| **Asthma** | 1,107,334 (11.5) | 27,864 (24.5) | 932,878 (11) | 61,132 (25) |
| **COPD** | 1,065,885 (11.1) | 19,837 (17.4) | 1,134,521 (13.4) | 47,793 (19.5) |
| **Sarcoidosis** | 10,515 (0.1) | 161 (0.1) | 11,423 (0.1) | 427 (0.2) |
| **RA** | 111,199 (1.2) | 1,399 (1.2) | 106,468 (1.3) | 2,930 (1.2) |
| **GERD** | 1,506,248 (15.7) | 16,849 (14.8) | 1,207,888 (14.2) | 33,245 (13.6) |
| **ARDS** | 20,652 (0.2) | 377 (0.3) | 37,815 (0.4) | 872 (0.4) |
| **IMV** | 196,580 (2) | 4,476 (3.9) | 240,354 (2.8) | 7,512 (3.1) |
| **Bacterial pneumonia** | 96,550 (1) | 3,516 (3.1) | 84,970 (1) | 6,032 (2.5) |
| **No PIPF** | 9,534,967 | 112,611 | 8,444,644 | 242,923 |
| **Incident PIPF** | 58,970 (0.6) | 1,236 (1.1) | 53,482 (0.6) | 1,826 (0.7) |
| **Attributable Risk** | 0.06 (-0.01, 0.13) | 4.86 (4.25, 5.47) | 0.03 (-0.04, 0.11) | 1.17 (0.82, 1.51) |
| **Attributable Fraction % (CI)** | 0.89 (0.78, 1.00) | 43.38 (40.12, 46.47) | 0.52 (0.36, 0.67) | 13.56 (9.32, 17.59) |
| **Risk Ratio** | 1.77 (1.67, 1.87) |  | 1.16 (1.10, 1.21) |  |
| **Univariate HR** | 1.53 (1.45, 1.63) |  | 1.32 (1.26, 1.40) |  |
| **Adjusted HR** | 1.46 (1.38, 1.55) |  | 1.26 (1.20, 1.33) |  |

**Abbreviations:** ARDS, Acute respiratory distress syndrome; COPD, Chronic Obstructive Pulmonary Disease; HR, Hazard Ratio; GERD, Gastroesophageal Reflux Disease; IMV, Invasive Mechanical Ventilation; PIPF, Post Inflammatory Pulmonary Fibrosis; RA, Rheumatoid Arthritis; Viral P, Viral pneumonia;

**Figure:**
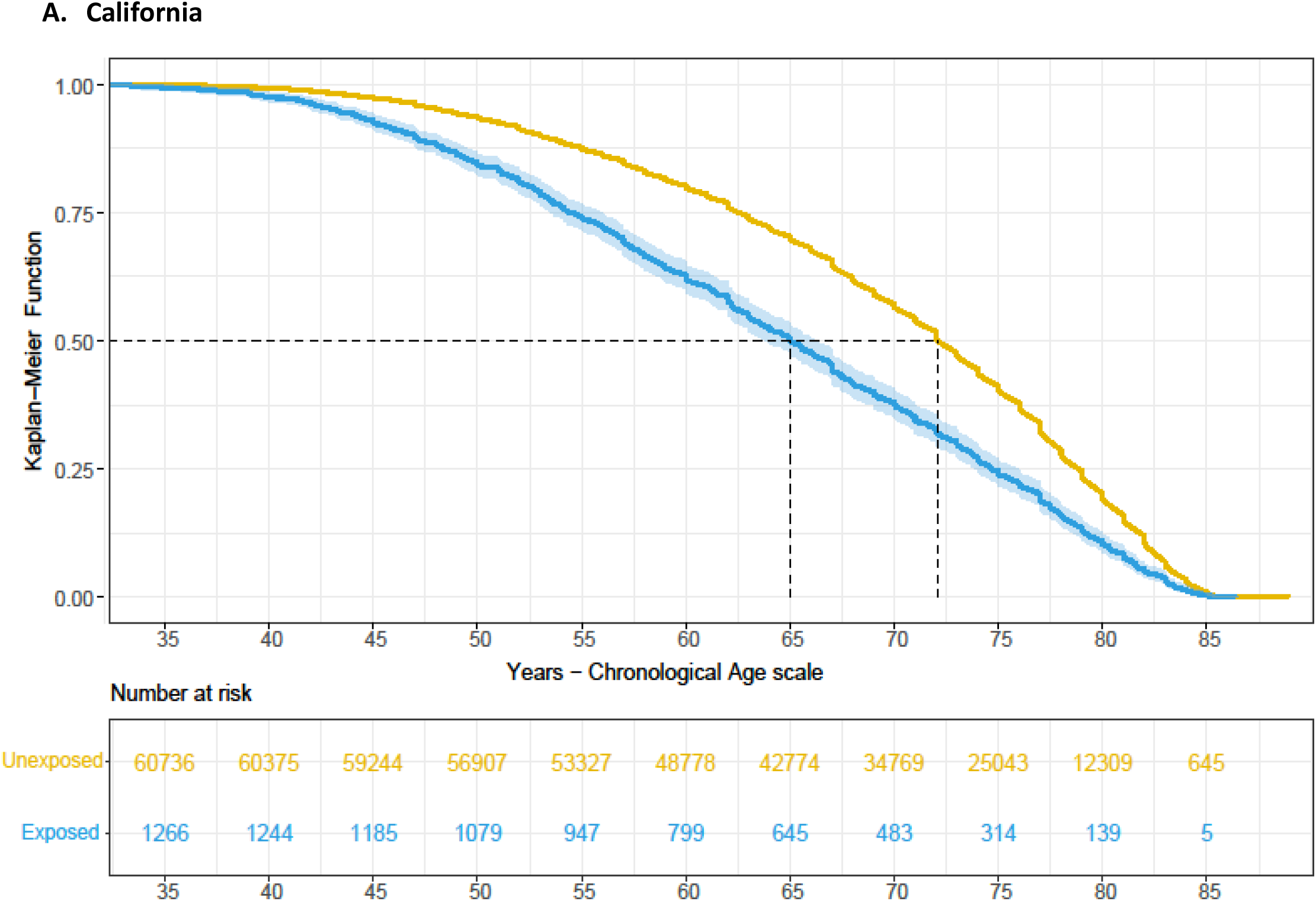
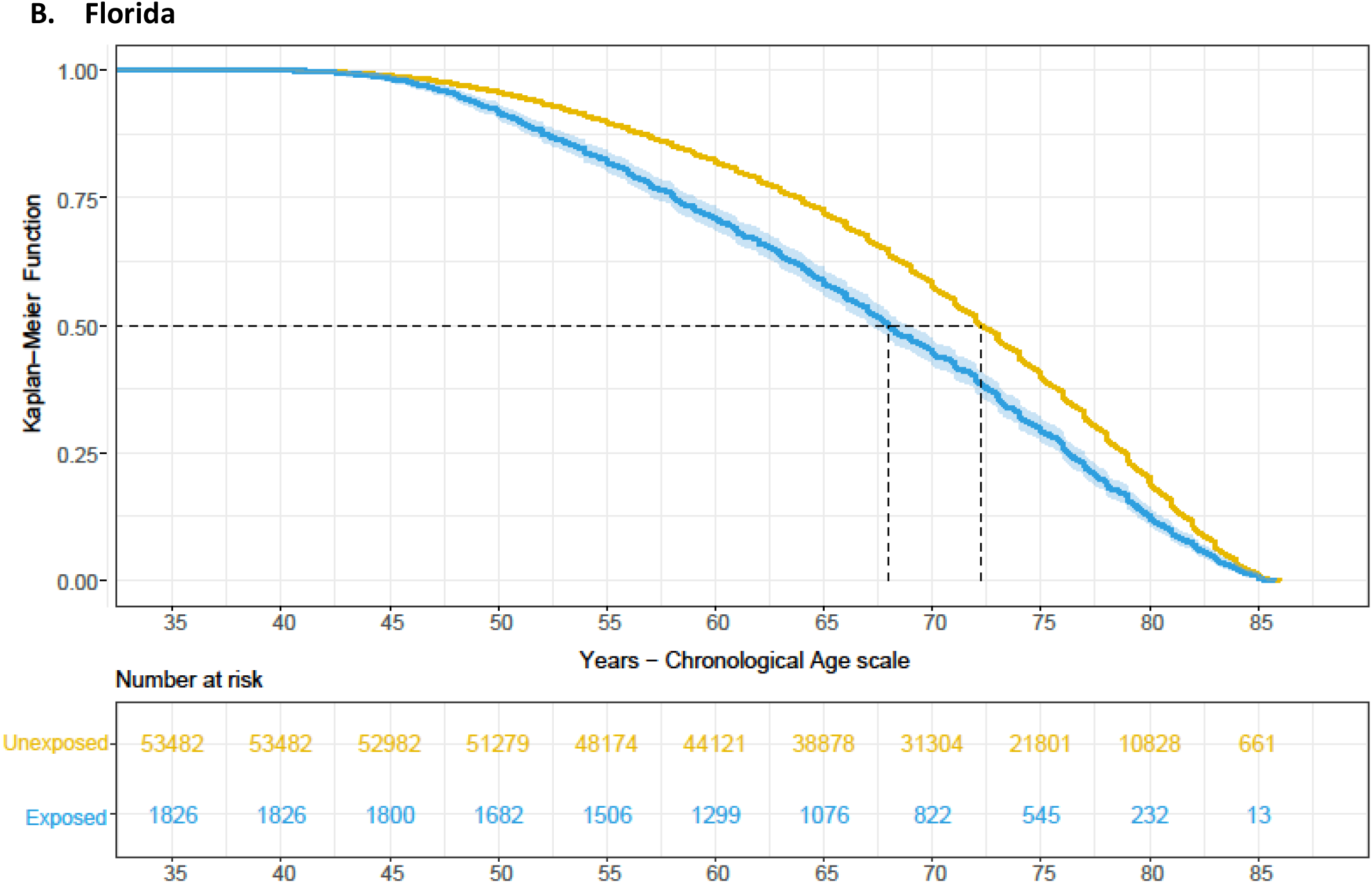
Kaplan Meier survival curves of all patients who had an incidence of Post Inflammatory Pulmonary Fibrosis (right censored and truncated) in California (A) and Florida (B) states. Patients exposed to Viral Pneumonia are shown in **blue** versus **others in brown**. Shaded regions represent 95% confidence intervals. Chronological age was used as the time scale (X-axis). The median time to diagnosis is 7.41 (6.16 −8.66) and 4.80 (4.34 − 5.26) earlier for patients with viral pneumonia versus without in California and Florida cohorts, respectively. The log-rank p <0.001 in both graphs.

## Discussion

Our results suggest that viral pneumonia is an independent risk factor for PIPF. The risk was increased by 46% and 26% in the discovery and validation cohorts, respectively. Moreover, among individuals eventually diagnosed with PIPF, those with viral pneumonia were younger and developed PIPF earlier. Limitations of the study include lack of information on disease severity and reliance on hospitalization ICD9 codes, which is likely to significantly underestimate viral pneumonia and PIPF diagnoses and identify PIPF patients at a later stage. Nevertheless, this is the first study demonstrating increased risk for post-inflammatory pulmonary fibrosis in patients with viral pneumonia in two large independent cohorts. Our findings suggest that patients hospitalized with viral pneumonia may have long term respiratory sequela that is often overlooked and suggest a need for additional studies focusing on phenotyping susceptible patients. This finding is especially important in light of the current COVID-19 pandemic because viral pneumonia is the most common manifestations of the disease (8), which could lead to subsequent fibrosis (9).

## Data Availability

The data is publicly available through Administrative health data from Healthcare Research and Quality (AHRQ) Healthcare Cost and Utilization Project (HCUP).

## References

1. HCUP State Emergency Department Databases (SEDD). Healthcare Cost and Utilization Project (HCUP). 2005-2015. Agency for Healthcare Research and Quality. Rockville, MD. www.hcup-us.ahrq.gov/sasdoverview.jsp.

2. HCUP State Inpatient Databases (SID). Healthcare Cost and Utilization Project (HCUP). 2005-2015. Agency for Healthcare Research and Quality. Rockville, MD. www.hcup-us.ahrq.gov/sasdoverview.jsp.

3. Houchens R. Inferences with HCUP State Databases Final Report: U.S. Agency for Healthcare Research and Quality; [updated October 12, 20102018/02/03]. Available from: http://www.hcup-us.ahrq.gov/reports/methods/methods.jsp.

4. Fingar KR OP, Barrett ML, Steiner CA. Using the HCUP Databases to Study Incidence and Prevalence: U.S. Agency for Healthcare Research and Quality; [updated December 6, 20162018/02/03]. Available from: http://www.hcup-us.ahrq.gov/reports/methods/methods.jsp.

5. Metcalfe D, Zogg CK, Haut ER, Pawlik TM, Haider AH, Perry DC. Data resource profile: State Inpatient Databases. Int J Epidemiol. 2019;48(6):1742–h.

6. Chalise P, Chicken E, McGee D. Time scales in epidemiological analysis: an empirical comparison. arXiv preprint arXiv:150202534. 2015.

7. Esposito DB, Lanes S, Donneyong M, Holick CN, Lasky JA, Lederer D, et al. Idiopathic pulmonary fibrosis in United States automated claims. Incidence, prevalence, and algorithm validation. American journal of respiratory and critical care medicine. 2015;192(10):1200–7.

8. Wu D, Wu T, Liu Q, Yang Z. The SARS-CoV-2 outbreak: What we know. International Journal of Infectious Diseases. 2020;94:44–8.

9. Spagnolo P, Balestro E, Aliberti S, Cocconcelli E, Biondini D, Della Casa G, et al. Pulmonary fibrosis secondary to COVID-19: a call to arms? The Lancet Respiratory Medicine. 2020.

